# Acceptability of Hepatitis C screening and treatment during pregnancy in pregnant women in Egypt, Pakistan and Ukraine

**DOI:** 10.1101/2021.09.29.21264171

**Authors:** Karen Scott, Elizabeth Chappell, Aya Mostafa, Alla Volokha, Nida Najmi, Fatma Ebeid, Svitlana Posokhova, Raheel Sikandar, Marta Vasylyev, Saima Zulfiqar, Viacheslav Kaminskyi, Sarah Pett, Ruslan Malyuta, Ruslana Karpus, Yomna Ayman, Rania H M Ahmed, Saeed Hamid, Manal H El-Sayed, Diana Gibb, Ali Judd, Intira Jeannie Collins

## Abstract

The risk of vertical transmission of hepatitis C virus (HCV) is ≈6%, and there is growing evidence that maternal HCV adversely affects pregnancy and infant outcomes. However, antenatal HCV screening is not routinely provided in most settings, and direct acting antivirals (DAA) are not approved for pregnant/ breastfeeding women. We conducted a cross-sectional survey of pregnant/post-partum women in Egypt, Pakistan and Ukraine to assess the acceptability of universal antenatal HCV screening and DAA treatment in the scenario of DAAs being approved for use in pregnancy. Among 630 women (n=210 per country), 73% were pregnant and 27% postpartum, 27% ever HCV antibody or PCR positive. Overall, 93% of women supported HCV screening and 88% would take DAAs in pregnancy (92%, 98% and 73% in Egypt, Pakistan and Ukraine, respectively), mostly to prevent vertical transmission/adverse pregnancy outcomes. Clinical trials to evaluate the safety and efficacy of DAAs in pregnancy are urgently needed.

## INTRODUCTION

Chronic hepatitis C (HCV) in women of childbearing age is a major global public health concern with an estimated 15 million women aged 15–49 years,^1^ living with HCV globally. The risk of HCV vertical transmission is ∼6%, with most of the estimated global population of 3 million children^2^ living with HCV thought to have acquired it this way. In addition there is growing evidence of adverse pregnancy and infant outcomes associated with maternal HCV, including intrahepatic cholestasis of pregnancy, preterm birth, and low birth weight.^3^

In 2016 the World Health Organization (WHO) adopted new goals for global elimination of viral hepatitis by 2030. However, pregnant and breastfeeding women and young children are currently excluded from HCV elimination strategies; few countries recommend routine screening in pregnancy, with many imposing additional charges which limits uptake,^4^ and direct acting antivirals (DAAs) are not licensed for use in pregnancy, breastfeeding or for children aged <3 years. International guidelines currently recommend pregnant women with chronic HCV (i.e. RNA positive) be referred for treatment after cessation of breastfeeding. However, in many low and middle-income countries (LMIC), women commonly breastfeed for ≥2 years, and have multiple pregnancies in rapid succession, limiting opportunities to treat. Studies in both high-income and LMIC have reported poor (<50%) retention in the HCV cascade of care among pregnant women and exposed infants, and low DAA uptake among women post-partum.^5, 6^

Antenatal HCV screening and treatment in pregnancy could cure maternal HCV and prevent vertical transmission and associated adverse pregnancy and infant outcomes, and substantially advance progress towards global elimination targets. A small phase I study in the USA assessed the pharmacokinetics of Harvoni® (sofosbuvir/ ledipasvir) in eight pregnant women from 24 weeks gestation; all women completed the 12-week treatment and had adequate drug exposure, all had viral suppression and functional cure by time of delivery, and no safety signals were identified.^7^ Other small studies have reported off-label use of DAAs in pregnant women in routine care, or conception during DAA treatment, again with no safety signals reported.^8, 9^ There is increasing recognition of the need for large clinical trials to assess the safety and efficacy of DAAs to cure pregnant women and prevent vertical transmission,^10^ as well as studies on likely uptake of HCV screening and treatment in pregnancy. One study, by Kushner et al, examined the acceptability of HCV screening and treatment in pregnancy in the USA. The study surveyed 121 non-pregnant women with current or previous chronic HCV infection; 60% reported willingness to take DAA treatment in pregnancy to reduce the risk of vertical transmission, and 21% would take treatment during pregnancy for maternal cure, even if it did not reduce vertical transmission.^11^ There are no comparable data from LMICs.

We conducted a cross-sectional survey to assess the acceptability of free universal antenatal HCV screening and potential uptake of DAAs in the scenario of DAAs being approved for use in pregnancy. The survey was conducted in three high HCV burden countries: Egypt, Pakistan and Ukraine. Egypt and Pakistan have generalised HCV epidemics with estimated prevalence in pregnant women of 9%^12^ and 6%^13^ respectively. Ukraine has a concentrated HCV epidemic with high prevalence among people living with HIV.^14^

## METHODS

Women aged ≥18 years who were pregnant or had delivered in the last 6 months, and attending participating antenatal clinics and maternity hospitals in Egypt (1 public facility), Pakistan (7 facilities, public and private) or Ukraine (3 public facilities, including 1 HIV antenatal clinic) were invited to participate. All women gave written informed consent. Ethical approval was obtained from ethics committees in each participating country.

A survey was developed in REDCap (Research Electronic Data Capture, www.project-redcap.org) and translated into local languages. Participants in Ukraine could self-complete the survey while those in Pakistan and Egypt completed it with assistance of study staff, due to high prevalence of poor literacy. Minor local adaptions were made, for example HIV status was not collected in Egypt or Pakistan as this was considered highly sensitive and both countries have low HIV prevalence and no routine HIV testing in pregnancy.^15, 16^ The survey ran from July 2020 to July 2021.

The first section of the survey collected data on socio-demographics, pregnancy, and HCV history and knowledge (24 questions about HCV, risk of transmission, availability of treatment and HCV in pregnancy, summarised as a composite). Participants were then given a factsheet about HCV and pregnancy, adapted from Kushner et al^11^ (Appendix 1). After they had read this, the second section collected views on the acceptability of free universal HCV screening and use of DAAs in pregnancy, in the scenario that participants had chronic HCV infection and that DAAs were safe and approved for use in pregnancy. The question about taking DAAs in pregnancy had three response options, adapted from Kushner et al.:^11^ (i) “Yes, to cure me, even if it does not lower the chance of my baby getting hepatitis C,”; (ii) “Yes, but only if it lowers the chance of my baby getting hepatitis C or has other potential benefits for my baby”; or (iii) “No, I would wait until after my pregnancy to start treatment.”

The target sample size was 630 women (210 per country), of which 50% in Ukraine would be HCV positive (self-reported ever PCR or antibody positive), and up to 20% in Egypt and Pakistan. Ukraine had a higher target as HCV screening is provided routinely to women living with HIV.

Descriptive statistics were used to summarise the characteristics of participants and survey response, overall and by country. Acceptability of treatment in pregnancy (yes for any reason vs. no) was described by key demographic and clinical characteristics: age, highest level of education, household overcrowding score (number of rooms excluding kitchen and bathroom/number of people in home), relationship status, employment status, pregnancy status (currently pregnant vs. given birth in the last 6 months), self-reported HCV status (ever HCV antibody or RNA positive, HCV negative and never tested) and HIV status (Ukraine only), number of previous children, and the composite HCV knowledge score.

## RESULTS

Sociodemographic characteristics of the 630 participants are shown in Table 1. Median age was slightly higher in Ukraine, at 33 [interquartile range 28,36] years, and lowest in Pakistan at 29 [26,33] years. Fifty-nine per cent of women in Ukraine self-reported as ever HCV positive compared to 3% and 20% in Egypt and Pakistan, respectively. A quarter of women in Egypt never had an HCV test compared to 10% in Pakistan and 7% in Ukraine. Forty percent of women in Ukraine were living with HIV, of whom 33% were ever HCV positive.

**Table 1.**
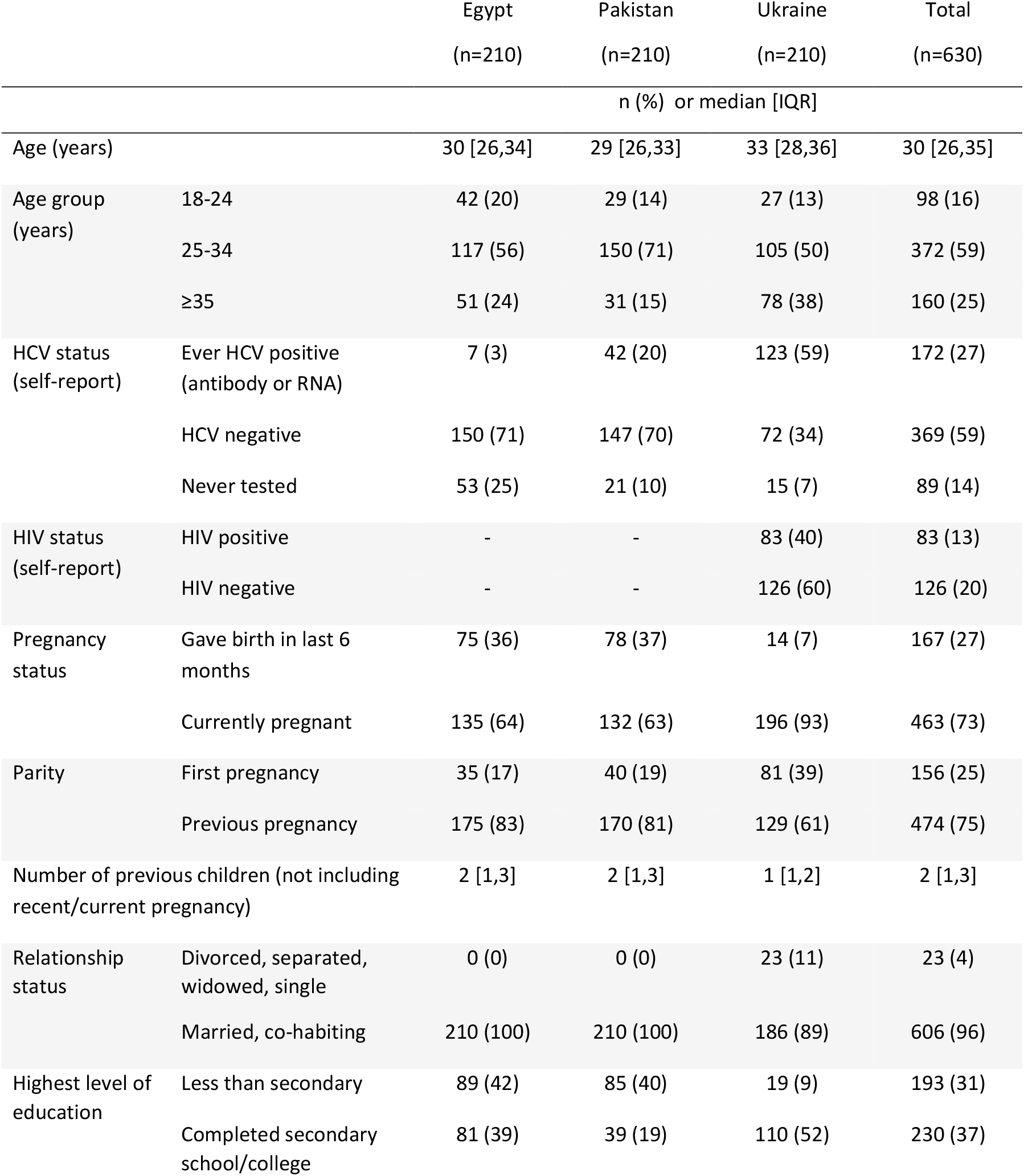
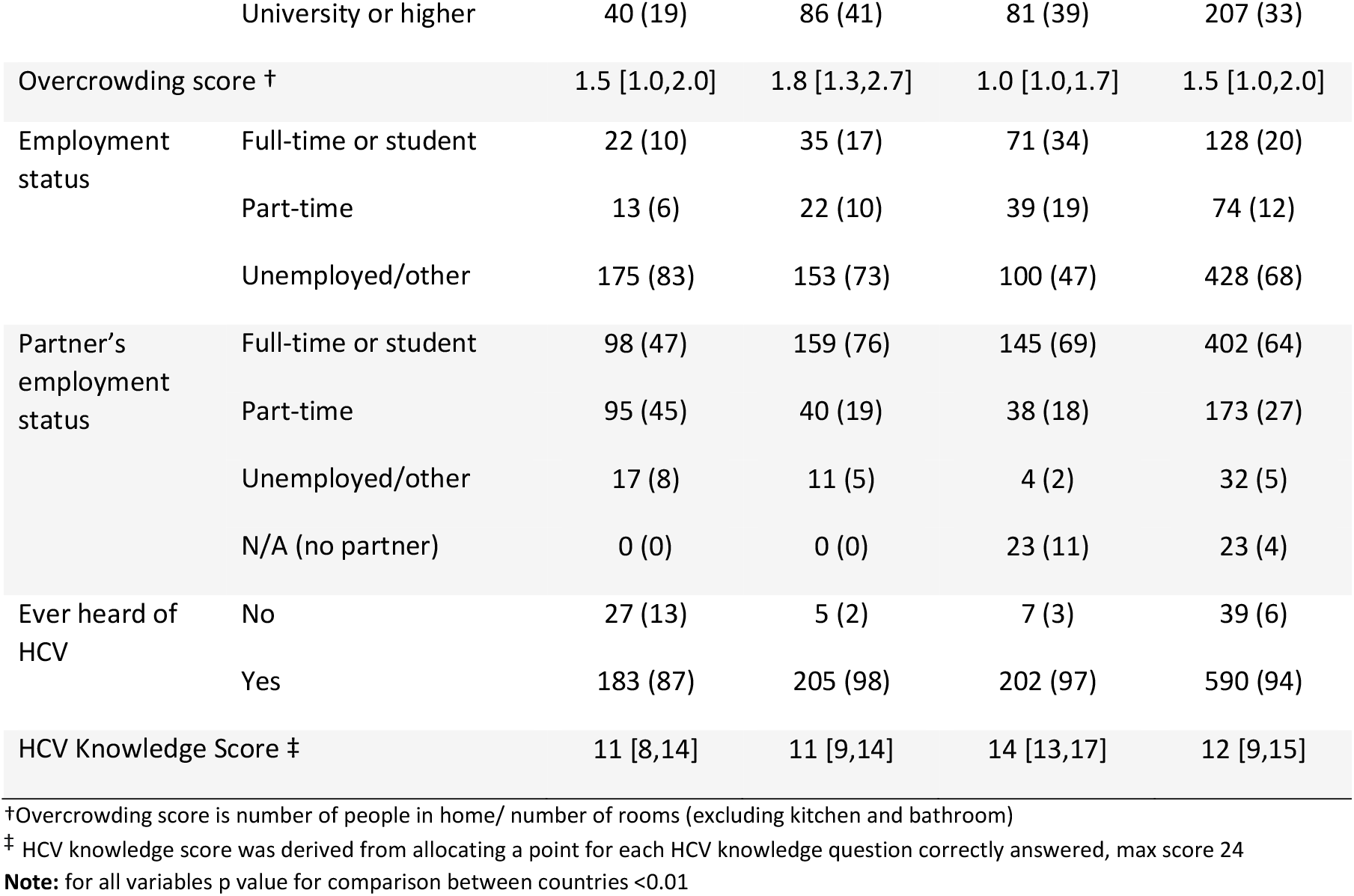
Sociodemographic characteristics of women participating in the survey by country

|  |  | Egypt<br>(n=210) | Pakistan<br>(n=210) | Ukraine<br>(n=210) | Total<br>(n=630) |
| --- | --- | --- | --- | --- | --- |
| n (%) or median [IQR] |  |  |  |  |  |
| Age (years) |  | 30 [26,34] | 29 [26,33] | 33 [28,36] | 30 [26,35] |
| Age group<br>(years) | 18-24 | 42 (20) | 29 (14) | 27 (13) | 98 (16) |
|  | 25-34 | 117 (56) | 150 (71) | 105 (50) | 372 (59) |
|  | ≥35 | 51 (24) | 31 (15) | 78 (38) | 160 (25) |
| HCV status<br>(self-report) | Ever HCV positive<br>(antibody or RNA) | 7 (3) | 42 (20) | 123 (59) | 172 (27) |
|  | HCV negative | 150 (71) | 147 (70) | 72 (34) | 369 (59) |
|  | Never tested | 53 (25) | 21 (10) | 15 (7) | 89 (14) |
| HIV status<br>(self-report) | HIV positive | - | - | 83 (40) | 83 (13) |
|  | HIV negative | - | - | 126 (60) | 126 (20) |
| Pregnancy<br>status | Gave birth in last 6<br>months | 75 (36) | 78 (37) | 14 (7) | 167 (27) |
|  | Currently pregnant | 135 (64) | 132 (63) | 196 (93) | 463 (73) |
| Parity | First pregnancy | 35 (17) | 40 (19) | 81 (39) | 156 (25) |
|  | Previous pregnancy | 175 (83) | 170 (81) | 129 (61) | 474 (75) |
| Number of previous children (not including<br>recent/current pregnancy) |  | 2 [1,3] | 2 [1,3] | 1 [1,2] | 2 [1,3] |
| Relationship<br>status | Divorced, separated,<br>widowed, single | 0 (0) | 0 (0) | 23 (11) | 23 (4) |
|  | Married, co-habiting | 210 (100) | 210 (100) | 186 (89) | 606 (96) |
| Highest level of<br>education | Less than secondary | 89 (42) | 85 (40) | 19 (9) | 193 (31) |
|  | Completed secondary<br>school/college | 81 (39) | 39 (19) | 110 (52) | 230 (37) |
|  | University or higher | 40 (19) | 86 (41) | 81 (39) | 207 (33) |
| Overcrowding score † |  | 1.5 [1.0,2.0] | 1.8 [1.3,2.7] | 1.0 [1.0,1.7] | 1.5 [1.0,2.0] |
| Employment status | Full-time or student | 22 (10) | 35 (17) | 71 (34) | 128 (20) |
|  | Part-time | 13 (6) | 22 (10) | 39 (19) | 74 (12) |
|  | Unemployed/other | 175 (83) | 153 (73) | 100 (47) | 428 (68) |
| Partner's employment status | Full-time or student | 98 (47) | 159 (76) | 145 (69) | 402 (64) |
|  | Part-time | 95 (45) | 40 (19) | 38 (18) | 173 (27) |
|  | Unemployed/other | 17 (8) | 11 (5) | 4 (2) | 32 (5) |
|  | N/A (no partner) | 0 (0) | 0 (0) | 23 (11) | 23 (4) |
| Ever heard of HCV | No | 27 (13) | 5 (2) | 7 (3) | 39 (6) |
|  | Yes | 183 (87) | 205 (98) | 202 (97) | 590 (94) |
| HCV Knowledge Score ‡ |  | 11 [8,14] | 11 [9,14] | 14 [13,17] | 12 [9,15] |

†Overcrowding score is number of people in home/ number of rooms (excluding kitchen and bathroom)
‡ HCV knowledge score was derived from allocating a point for each HCV knowledge question correctly answered, max score 24
**Note:** for all variables p value for comparison between countries <0.01

The majority of women in Ukraine (93%) were pregnant at the time of survey compared to two-thirds in Egypt and Pakistan. Most women were married or co-habiting. In Egypt and Pakistan ∼40% of women had not completed secondary school education, compared to 9% in Ukraine. The household overcrowding score was highest in Pakistan, signifying more overcrowding. The median HCV knowledge score was 12 [9,15] (of a possible maximum 24) and was comparable across countries, however 13% of women in Egypt reported not having heard of HCV before, compared to ≤3% in Pakistan and Ukraine.

Overall, 93% of women agreed with free universal HCV screening during pregnancy and this was consistent across countries (range 88%-95%) (Supplementary Figure 1). The acceptability of DAAs in pregnancy by country, HCV status, and HIV status (Ukraine only) is shown in Figure 1. Overall 88% of women reported that they would take DAAs in pregnancy; acceptability was highest in Pakistan (98%) followed by Egypt (91%) and Ukraine (73%). The majority of women would take DAAs in pregnancy to prevent vertical transmission and other potential benefits to the baby (48-90% by country), while a smaller proportion (8-29%) would take DAAs for maternal cure. When results were combined across all countries and stratified by HCV status, acceptability was highest among HCV negative women (92%) and was lowest among women who ever tested HCV positive at 78%. Among women in Ukraine, acceptability was similar among women with and without HIV infection.

**Figure 1.**
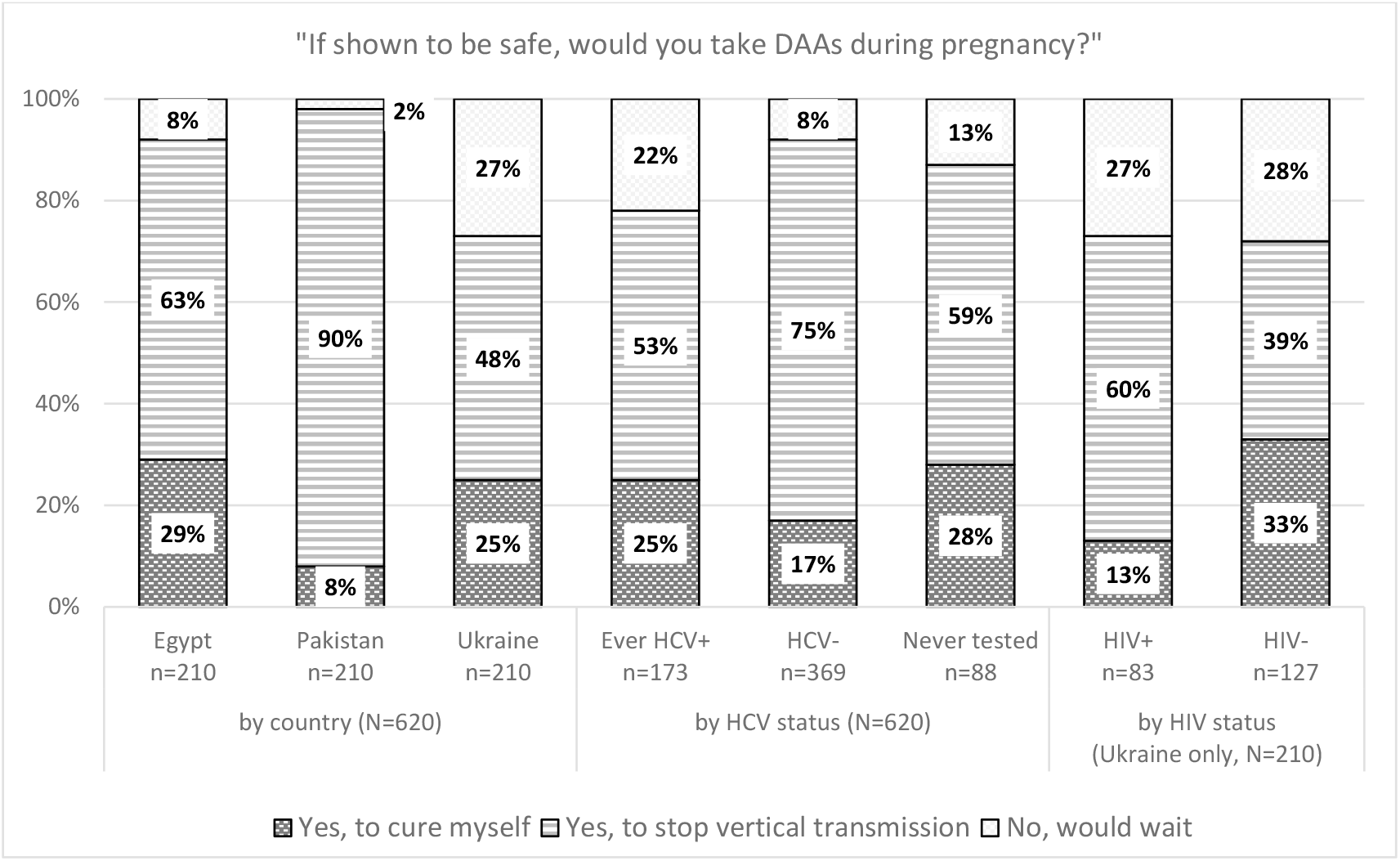
Acceptability of DAA treatment if known to be safe during pregnancy, by country, self-reported HCV status and self-reported HIV status (among women in Ukraine)

There was no difference in acceptability of DAAs by the woman’s or her partner’s employment status, or her HCV knowledge score, in any country (all p>0.05) (Supplementary Table 1). In Egypt, only higher household overcrowding was associated with increased acceptability (p=0.018). In Pakistan, older maternal age (p=0.007), fewer previous children (p=0.032), and higher household overcrowding (p=0.047) were associated with increased acceptability. In Ukraine, being pregnant (p=0.047) and higher education level (p=0.031) were associated with increased acceptability.

## DISCUSSION

To our knowledge, this is the largest study to date on acceptability of HCV screening and treatment in pregnant/post-partum women and first in LMICs with high HCV burden. Acceptability of universal antenatal HCV screening was very high at 93%. In the scenario that DAAs were approved for use in pregnancy, acceptability ranged from 78% in Ukraine to 98% in Pakistan. These estimates are substantially higher than the 60% acceptability of DAAs in pregnancy reported among non-pregnant women with current or past history of chronic HCV infection in the USA.^11^ The difference may be due to our study including women who were currently pregnant or recently delivered and women with known and unknown HCV status,. It may also reflect the broader differences in DAA acceptability during pregnancy across different settings.

In our study, the majority of women would take DAAs in pregnancy to prevent vertical transmission and adverse pregnancy and infant outcomes while fewer reported maternal cure as the main reason to receive treatment. This trend was consistent across the three countries, by HCV and HIV status. We found few individual level factors associated with the acceptability of DAAs, which was in part due the high acceptability observed.

Interestingly, despite a national HCV test and treat campaign in Egypt, with over 50 million screened and 4 million treated since 2015,^17^ a small proportion of women (13%) were not aware of HCV, and 25% were never tested. We gave all participants a factsheet to encourage informed responses, however we did not assess whether this information was fully understood or the degree to which it contributed to women’s responses.

One of the strengths of our study is the inclusion of women from three countries in different geographic regions with different HCV epidemics, and including public and private clinics and an HIV clinic. A key limitation is that assessment of acceptability was based on a scenario that the woman had chronic HCV, and that DAAs were safe and approved for use in pregnancy, yet the majority of women included in the study in Pakistan and Egypt were HCV negative or had never been tested. However almost 60% of the women in Ukraine were HCV positive, and for this group acceptability was high at 88%. Nonetheless, actual uptake of DAA treatments if approved may differ. Another weakness of the study is that the method of data collection differed across settings whereby most women in Ukraine self-completed the survey on a tablet and all women in Egypt and Pakistan were surveyed by a member of staff. This may have influenced women’s responses.

In summary our study found very high acceptability of universal HCV antenatal screening and the use of DAAs in pregnancy, if found to be safe and approved for use in pregnancy. Currently pregnant women with chronic HCV have to wait until they finish breastfeeding before they can seek curative treatment, resulting in long delays in access to treatment and increased risk of disease progression, which may affect maternal and infant outcomes for current and future pregnancies. Studies in the USA^18^ and France^19^ have reported the cost-effectiveness of universal HCV screening in pregnancy even without the option of treatment during pregnancy, driven by the benefits of identifying women to refer for treatment after breastfeeding and HCV exposed infants to screen. There are no comparable cost-effectiveness analyses in LMIC but preliminary modelling of universal screening and treatment in Ukraine and Egypt suggests substantial potential benefits.^20^ A DAA pregnancy registry, the Treatment In Pregnancy for Hepatitis C: TiP-HepC Registry, has been established by the Coalition for Global Hepatitis Elimination at the Taskforce for Global Health to aid reporting of first trimester exposure to DAAs, which will contribute important safety data. Additionally randomised controlled trials on the safety and efficacy of DAAs started later in pregnancy and the breastfeeding period are needed to inform future clinical care and policy to ensure that pregnant women with HCV and their children do not continue to be left behind in the global HCV elimination effort.

## Data Availability

Data sharing requests may be made in writing to UCL

## Abbreviations

HCV: Hepatitis C virus
DAAs: Direct acting antivirals
PCR: Polymerase chain reaction
WHO: World Health Organization
RNA: Ribonucleic acid
LMIC: Low and middle-income countries
HIV: Human immunodeficiency virus

## Supplementary Figures and Tables

**Supp. Figure 1:**
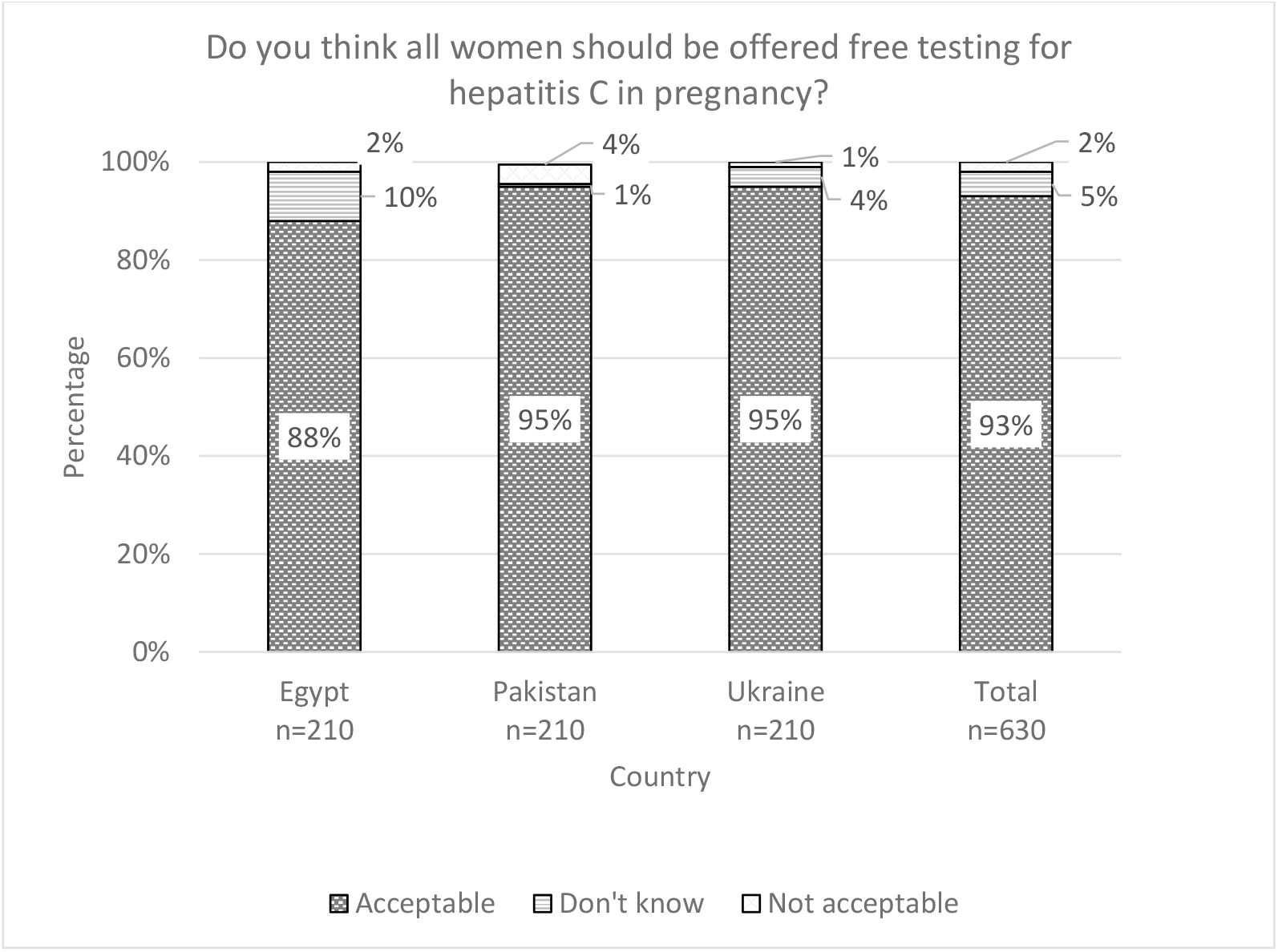
Acceptability of universal HCV screening in pregnancy

**Supp Table 1:**
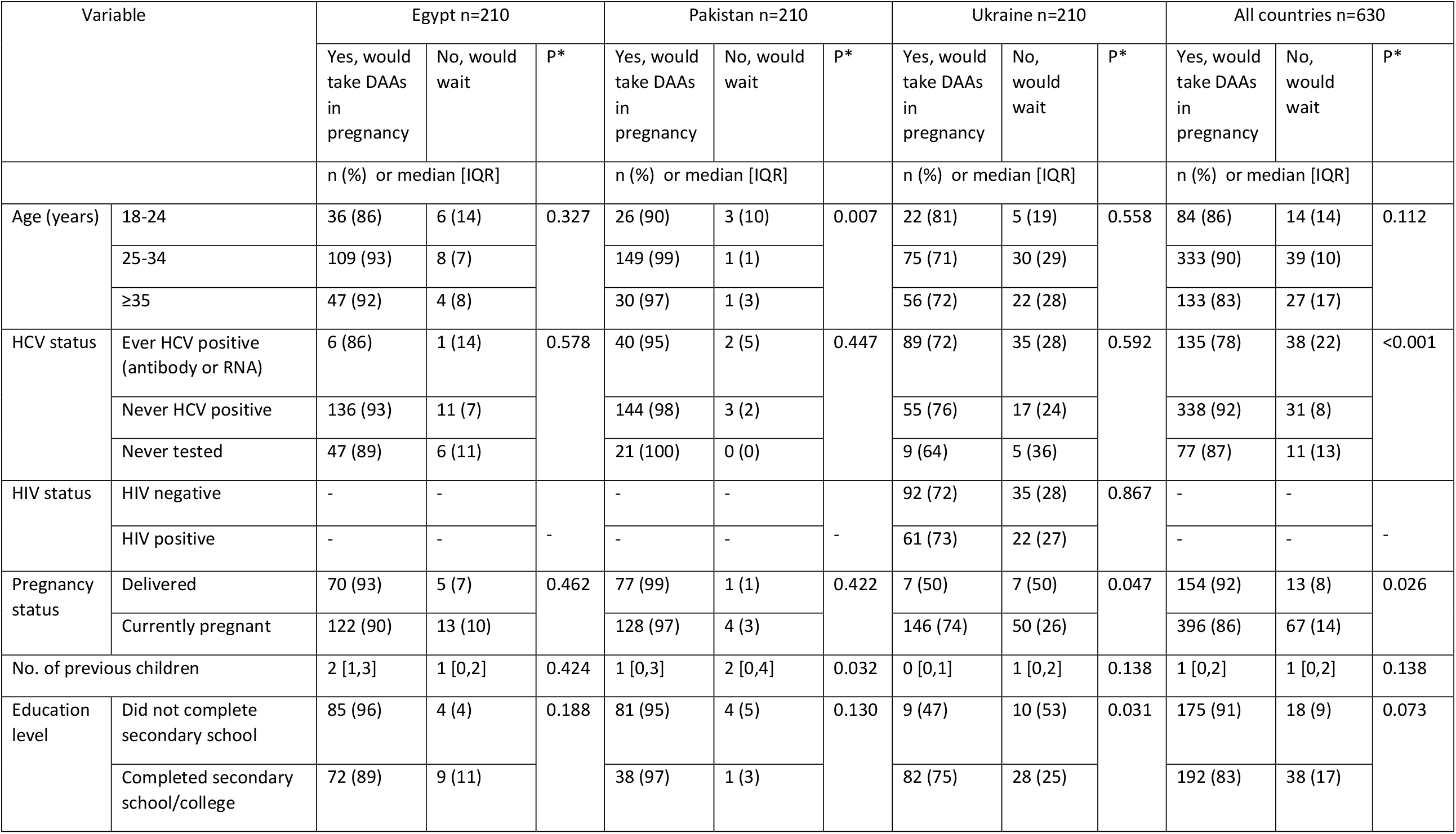
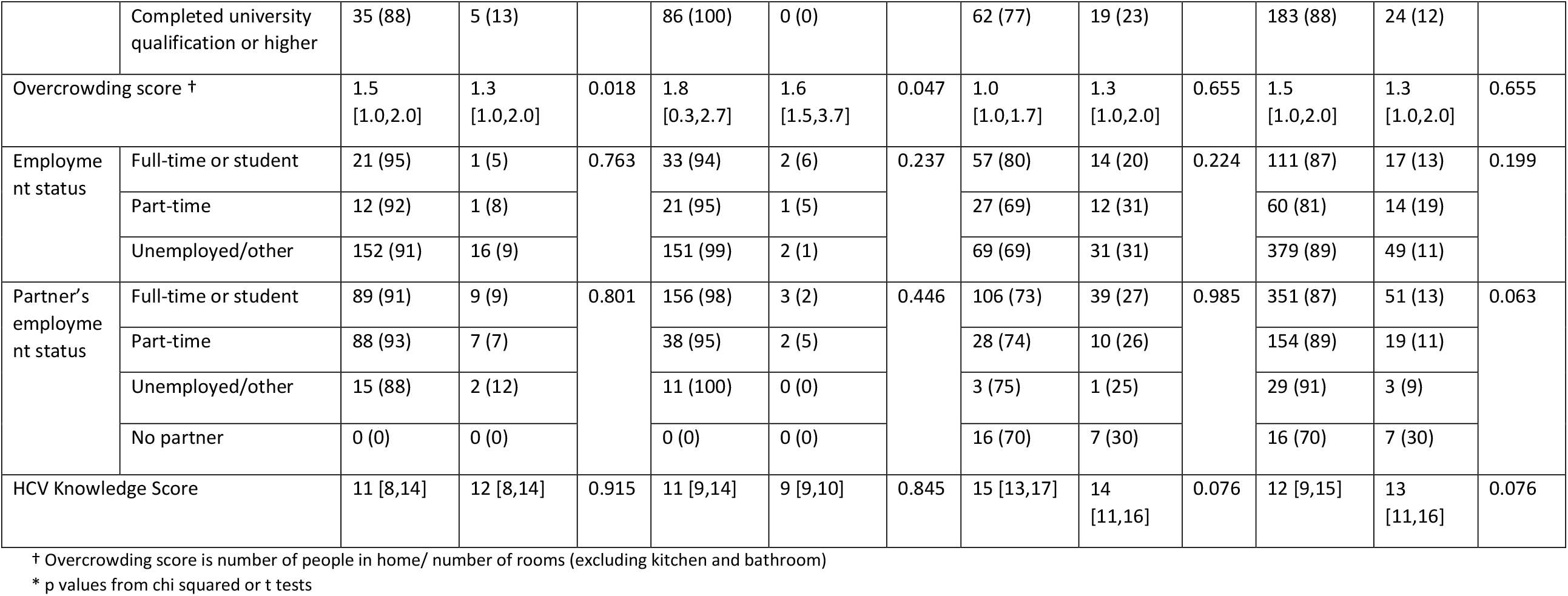
Acceptability of DAAs in pregnancy by key characteristics, by country and overall

### Appendix 1 HCVAVERT Screening and DAA Acceptability Survey for Ukraine

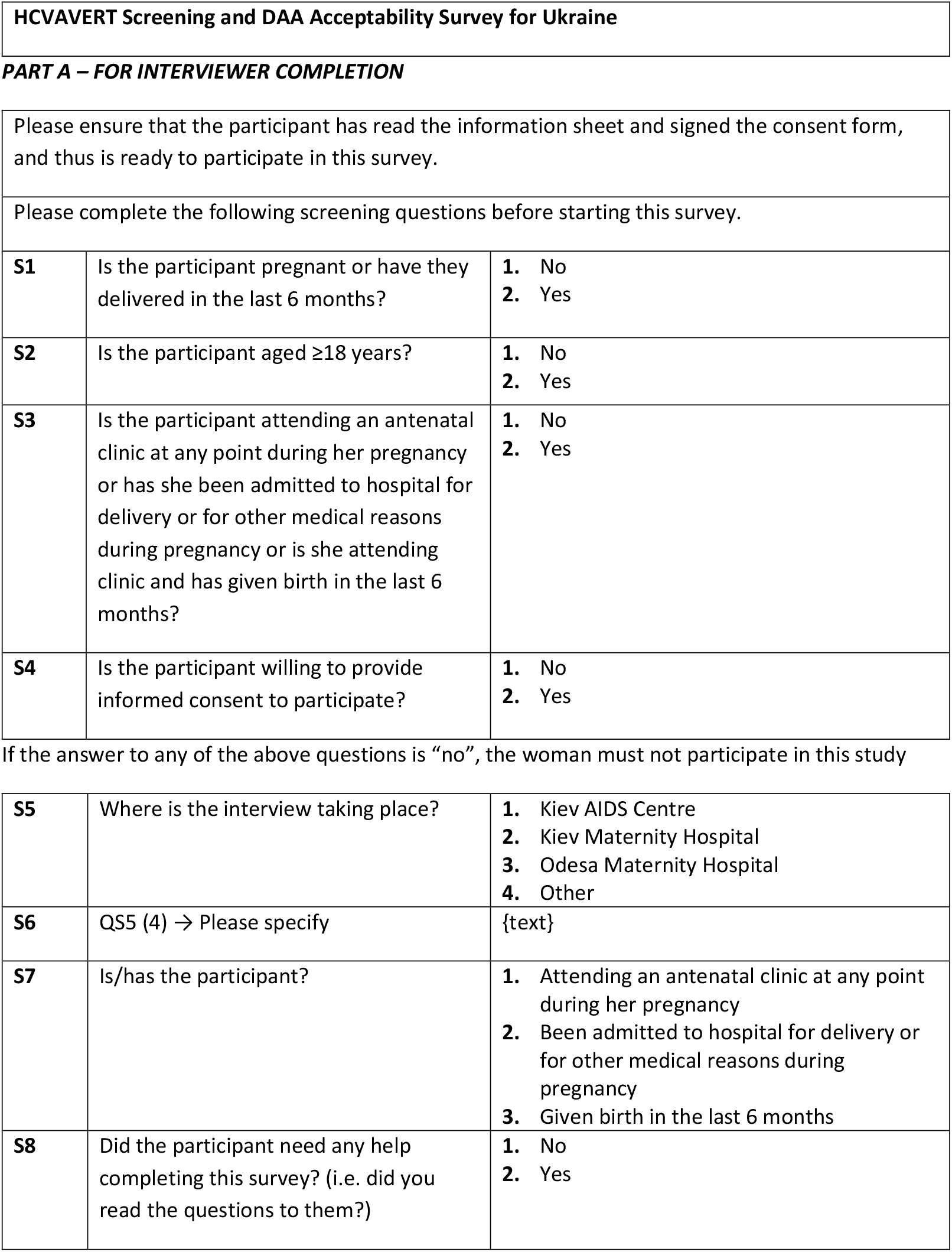
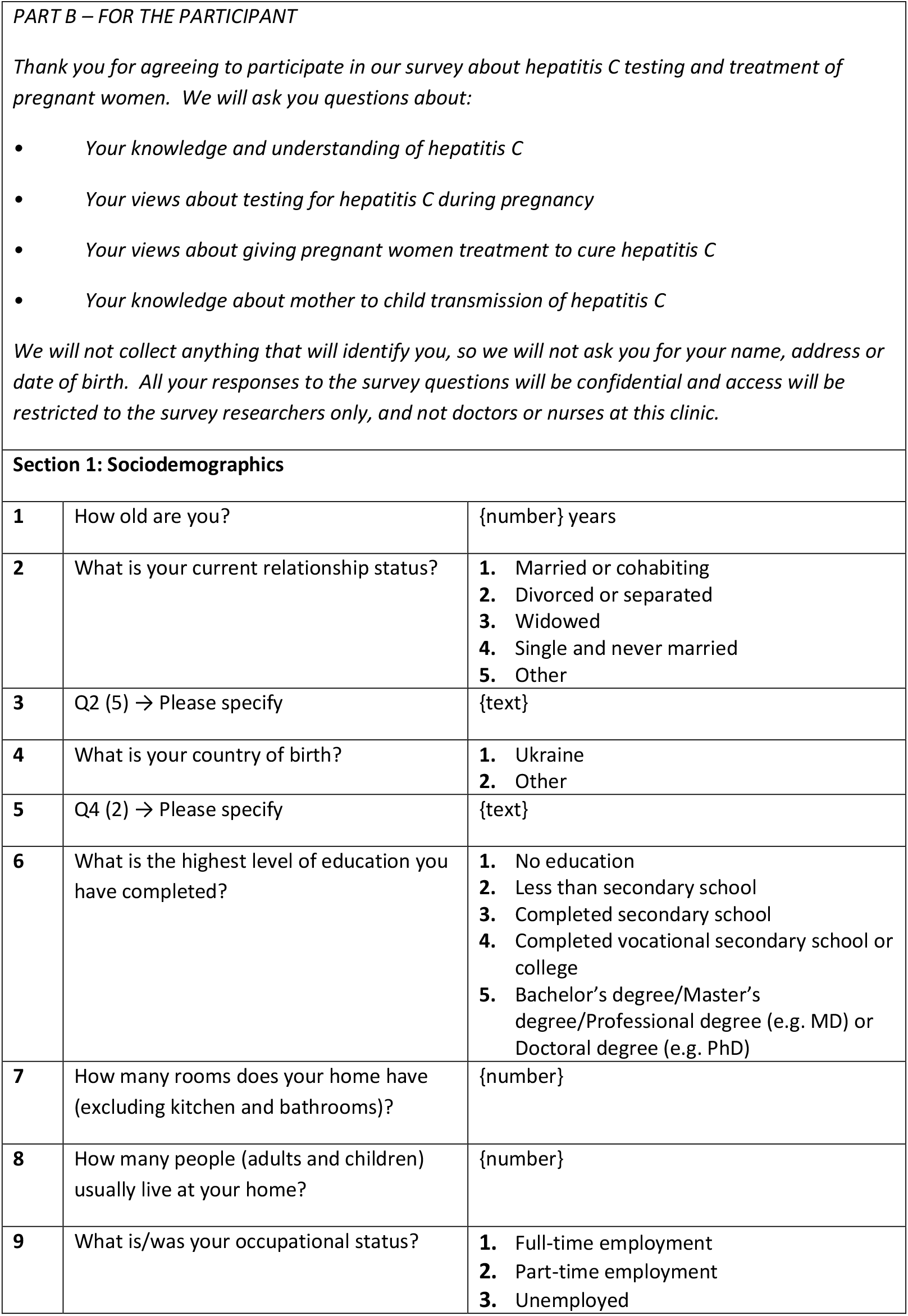
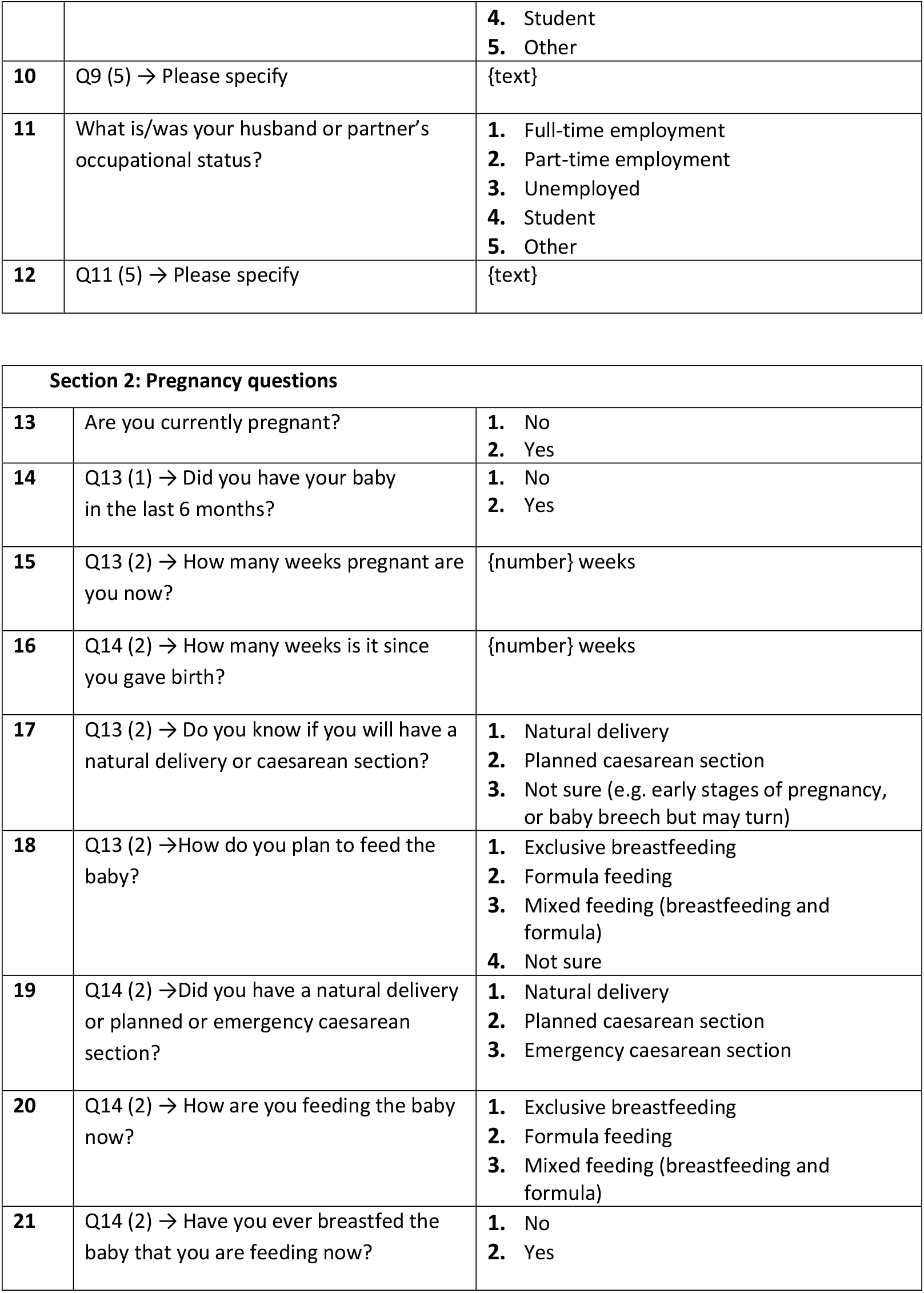
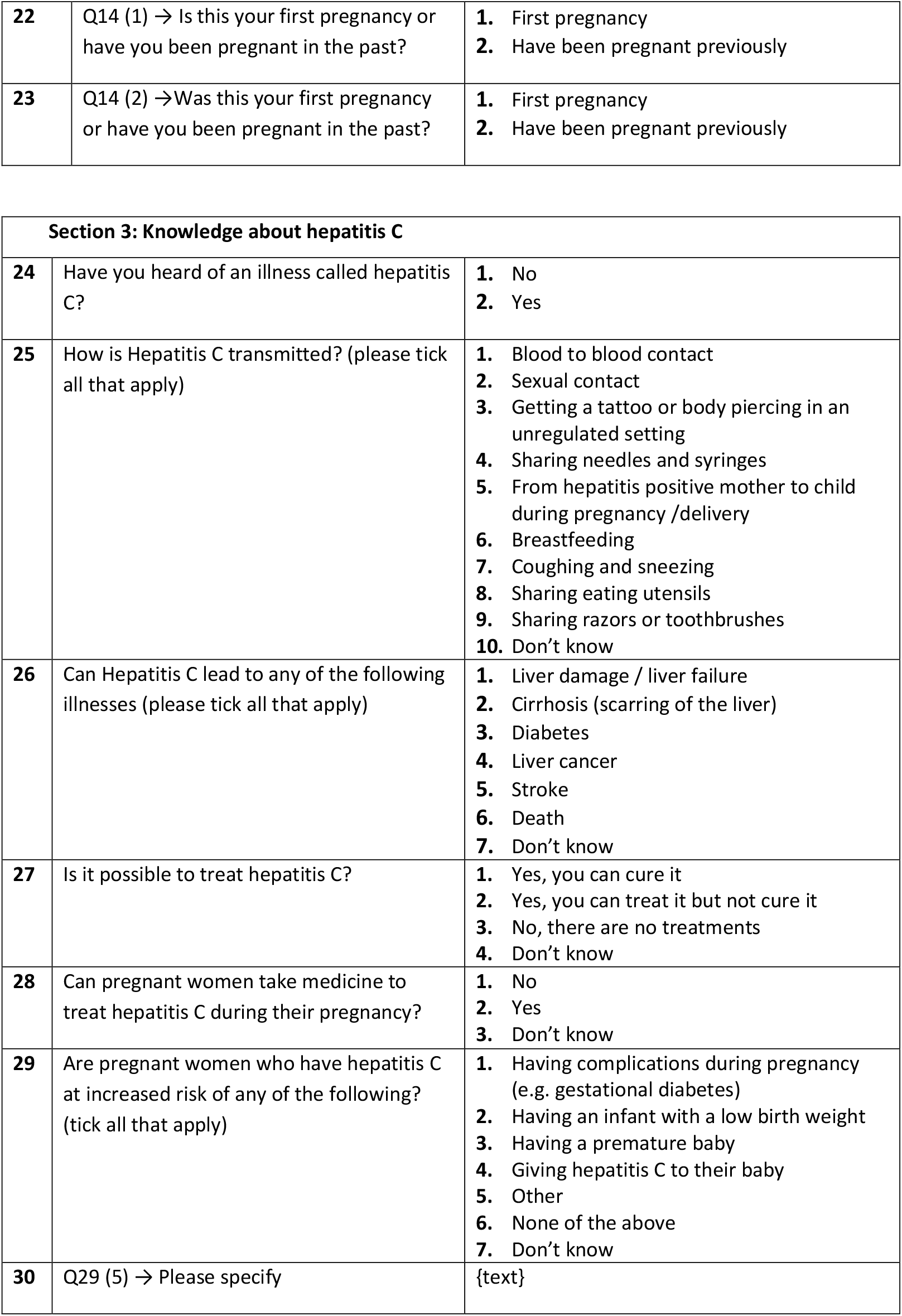
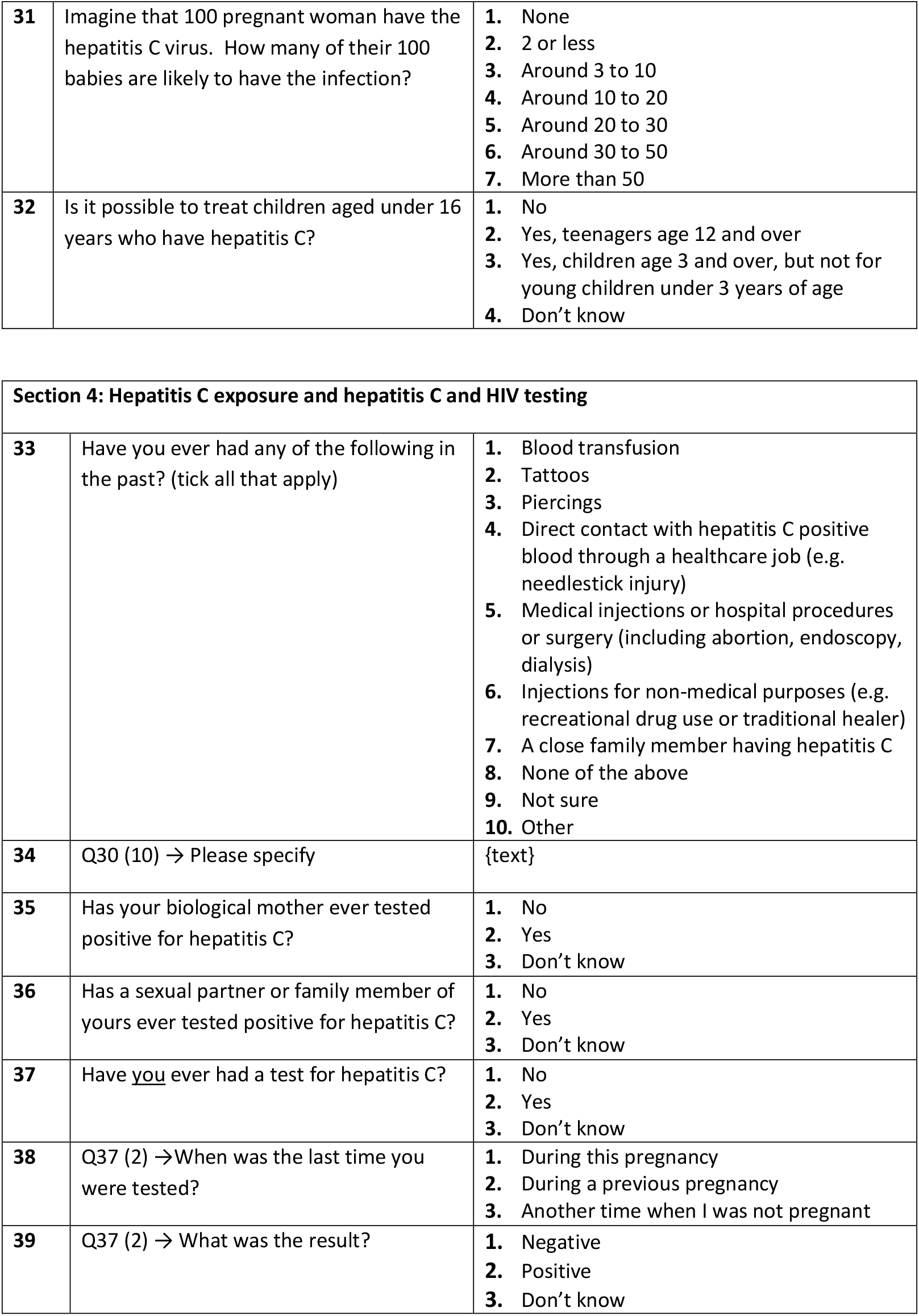
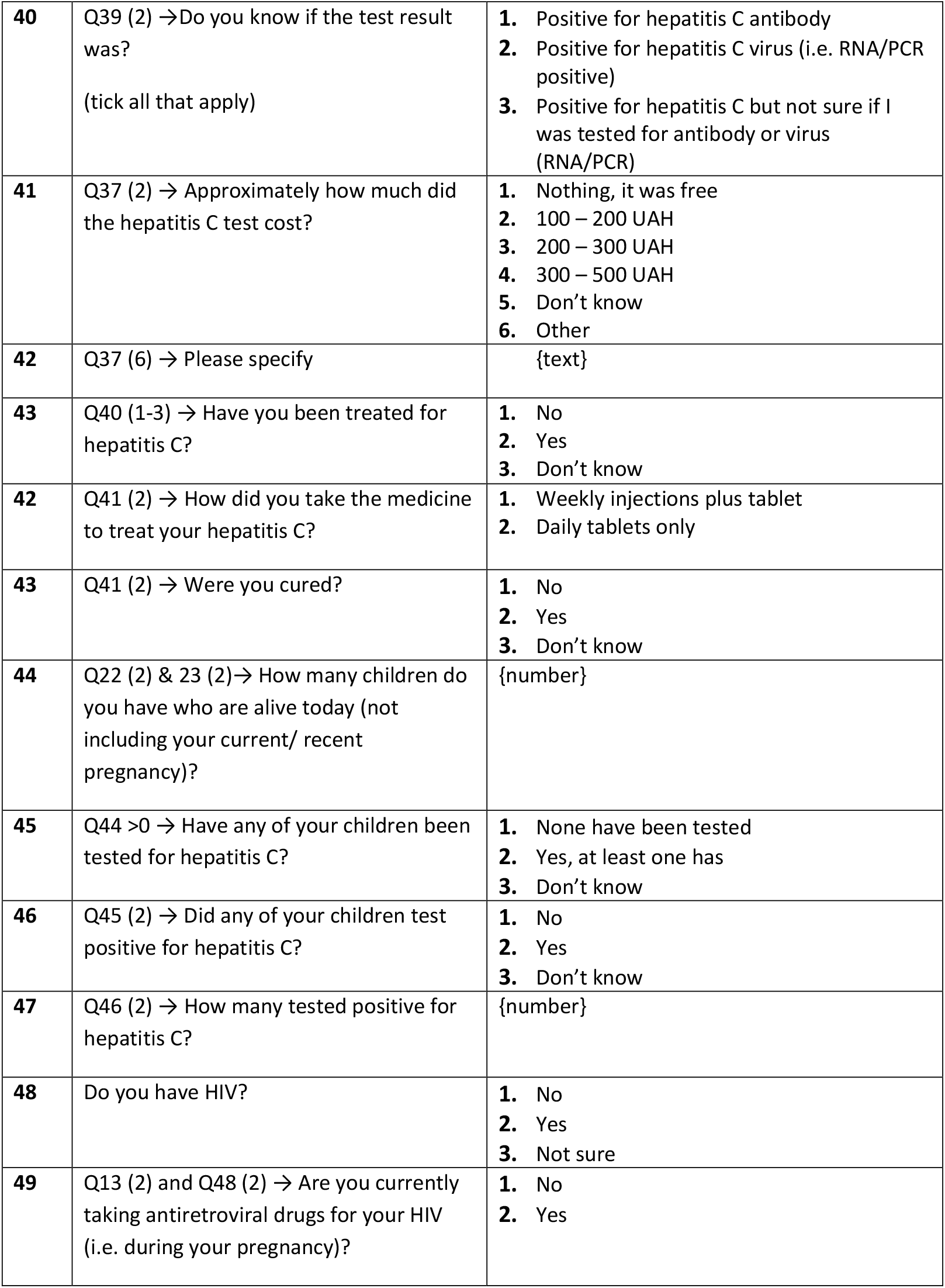
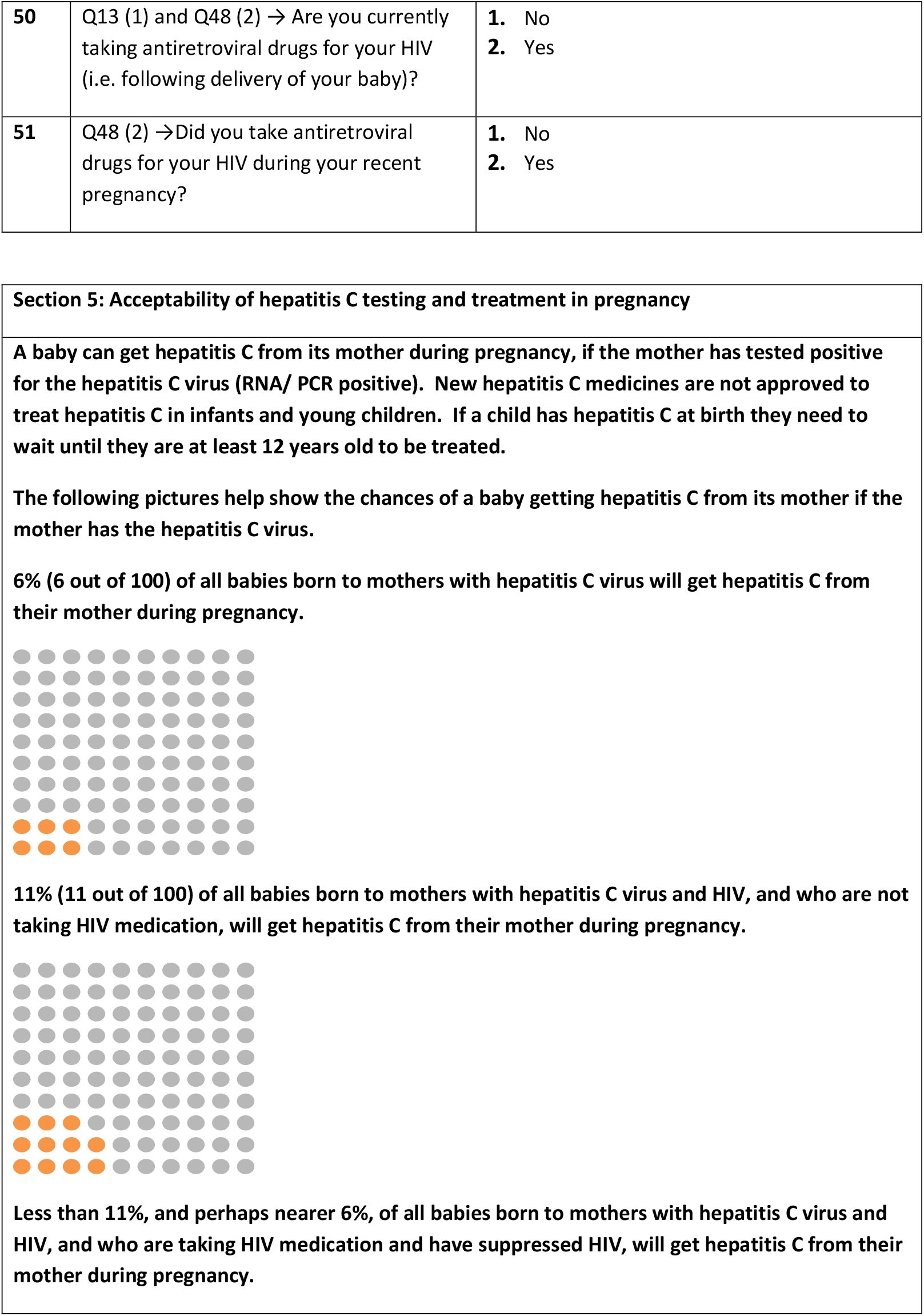
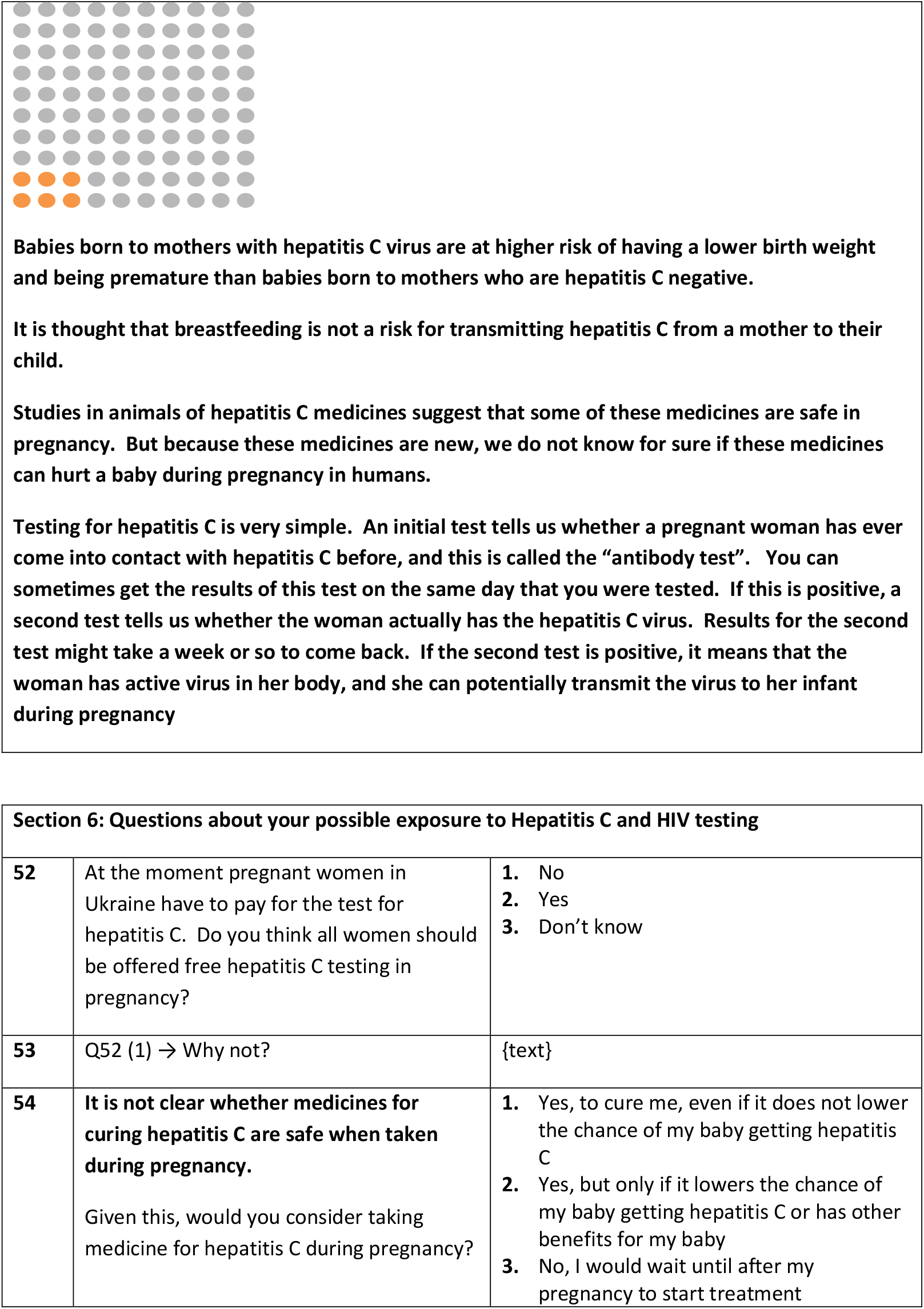
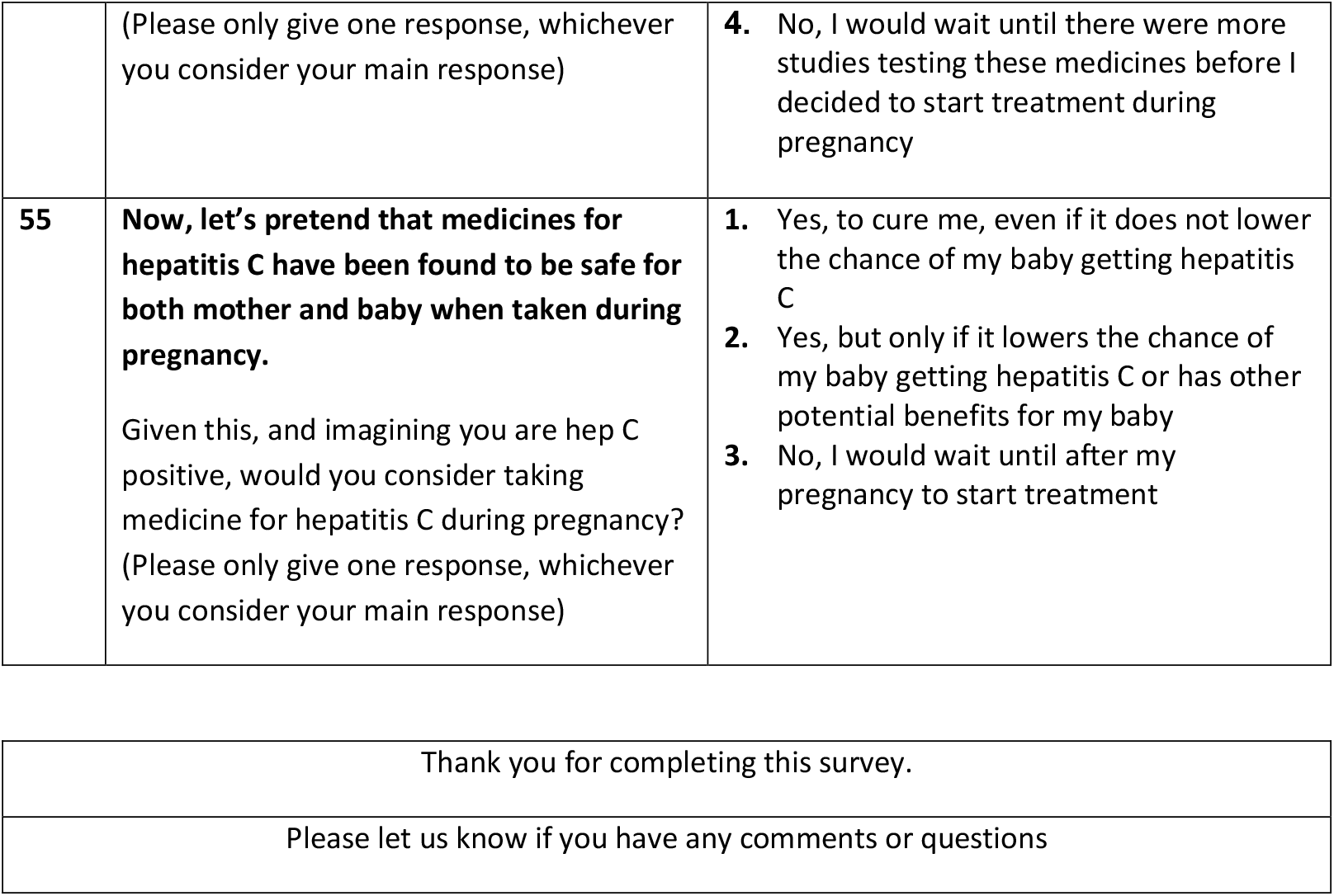

|  |  |  |
| --- | --- | --- |
| **S1** | Is the participant pregnant or have they delivered in the last 6 months? | 1. No<br>2. Yes |
| **S2** | Is the participant aged $\geq 18$ years? | 1. No<br>2. Yes |
| **S3** | Is the participant attending an antenatal clinic at any point during her pregnancy or has she been admitted to hospital for delivery or for other medical reasons during pregnancy or is she attending clinic and has given birth in the last 6 months? | 1. No<br>2. Yes |
| **S4** | Is the participant willing to provide informed consent to participate? | 1. No<br>2. Yes |
| If the answer to any of the above questions is “no”, the woman must not participate in this study |  |  |
| **S5** | Where is the interview taking place? | 1. Kiev AIDS Centre<br>2. Kiev Maternity Hospital<br>3. Odesa Maternity Hospital<br>4. Other |
| **S6** | QS5 (4) → Please specify | {text} |
| **S7** | Is/has the participant? | 1. Attending an antenatal clinic at any point during her pregnancy<br>2. Been admitted to hospital for delivery or for other medical reasons during pregnancy<br>3. Given birth in the last 6 months |
| **S8** | Did the participant need any help completing this survey? (i.e. did you read the questions to them?) | 1. No<br>2. Yes |
| **Section 1: Sociodemographics** |  |  |
| **1** | How old are you? | {number} years |
| **2** | What is your current relationship status? | **1.** Married or cohabiting<br>**2.** Divorced or separated<br>**3.** Widowed<br>**4.** Single and never married<br>**5.** Other |
| **3** | Q2 (5) → Please specify | {text} |
| **4** | What is your country of birth? | **1.** Ukraine<br>**2.** Other |
| **5** | Q4 (2) → Please specify | {text} |
| **6** | What is the highest level of education you have completed? | **1.** No education<br>**2.** Less than secondary school<br>**3.** Completed secondary school<br>**4.** Completed vocational secondary school or college<br>**5.** Bachelor's degree/Master's degree/Professional degree (e.g. MD) or Doctoral degree (e.g. PhD) |
| **7** | How many rooms does your home have (excluding kitchen and bathrooms)? | {number} |
| **8** | How many people (adults and children) usually live at your home? | {number} |
| **9** | What is/was your occupational status? | **1.** Full-time employment<br>**2.** Part-time employment<br>**3.** Unemployed |
|  |  | **4. Student**<br>**5. Other** |
| **10** | Q9 (5) → Please specify | {text} |
| **11** | What is/was your husband or partner's occupational status? | **1. Full-time employment**<br>**2. Part-time employment**<br>**3. Unemployed**<br>**4. Student**<br>**5. Other** |
| **12** | Q11 (5) → Please specify | {text} |

| **Section 2: Pregnancy questions** |  |  |
| --- | --- | --- |
| **13** | Are you currently pregnant? | **1. No**<br>**2. Yes** |
| **14** | Q13 (1) → Did you have your baby in the last 6 months? | **1. No**<br>**2. Yes** |
| **15** | Q13 (2) → How many weeks pregnant are you now? | {number} weeks |
| **16** | Q14 (2) → How many weeks is it since you gave birth? | {number} weeks |
| **17** | Q13 (2) → Do you know if you will have a natural delivery or caesarean section? | **1. Natural delivery**<br>**2. Planned caesarean section**<br>**3. Not sure (e.g. early stages of pregnancy, or baby breech but may turn)** |
| **18** | Q13 (2) → How do you plan to feed the baby? | **1. Exclusive breastfeeding**<br>**2. Formula feeding**<br>**3. Mixed feeding (breastfeeding and formula)**<br>**4. Not sure** |
| **19** | Q14 (2) → Did you have a natural delivery or planned or emergency caesarean section? | **1. Natural delivery**<br>**2. Planned caesarean section**<br>**3. Emergency caesarean section** |
| **20** | Q14 (2) → How are you feeding the baby now? | **1. Exclusive breastfeeding**<br>**2. Formula feeding**<br>**3. Mixed feeding (breastfeeding and formula)** |
| **21** | Q14 (2) → Have you ever breastfed the baby that you are feeding now? | **1. No**<br>**2. Yes** |
| **22** | Q14 (1) → Is this your first pregnancy or have you been pregnant in the past? | **1.** First pregnancy<br>**2.** Have been pregnant previously |
| **23** | Q14 (2) → Was this your first pregnancy or have you been pregnant in the past? | **1.** First pregnancy<br>**2.** Have been pregnant previously |

| **Section 3: Knowledge about hepatitis C** |  |  |
| --- | --- | --- |
| **24** | Have you heard of an illness called hepatitis C? | **1.** No<br>**2.** Yes |
| **25** | How is Hepatitis C transmitted? (please tick all that apply) | **1.** Blood to blood contact<br>**2.** Sexual contact<br>**3.** Getting a tattoo or body piercing in an unregulated setting<br>**4.** Sharing needles and syringes<br>**5.** From hepatitis positive mother to child during pregnancy /delivery<br>**6.** Breastfeeding<br>**7.** Coughing and sneezing<br>**8.** Sharing eating utensils<br>**9.** Sharing razors or toothbrushes<br>**10.** Don't know |
| **26** | Can Hepatitis C lead to any of the following illnesses (please tick all that apply) | **1.** Liver damage / liver failure<br>**2.** Cirrhosis (scarring of the liver)<br>**3.** Diabetes<br>**4.** Liver cancer<br>**5.** Stroke<br>**6.** Death<br>**7.** Don't know |
| **27** | Is it possible to treat hepatitis C? | **1.** Yes, you can cure it<br>**2.** Yes, you can treat it but not cure it<br>**3.** No, there are no treatments<br>**4.** Don't know |
| **28** | Can pregnant women take medicine to treat hepatitis C during their pregnancy? | **1.** No<br>**2.** Yes<br>**3.** Don't know |
| **29** | Are pregnant women who have hepatitis C at increased risk of any of the following? (tick all that apply) | **1.** Having complications during pregnancy (e.g. gestational diabetes)<br>**2.** Having an infant with a low birth weight<br>**3.** Having a premature baby<br>**4.** Giving hepatitis C to their baby<br>**5.** Other<br>**6.** None of the above<br>**7.** Don't know |
| **30** | Q29 (5) → Please specify | {text} |
| **31** | Imagine that 100 pregnant woman have the hepatitis C virus. How many of their 100 babies are likely to have the infection? | 1. None<br>2. 2 or less<br>3. Around 3 to 10<br>4. Around 10 to 20<br>5. Around 20 to 30<br>6. Around 30 to 50<br>7. More than 50 |
| **32** | Is it possible to treat children aged under 16 years who have hepatitis C? | 1. No<br>2. Yes, teenagers age 12 and over<br>3. Yes, children age 3 and over, but not for young children under 3 years of age<br>4. Don't know |

| **Section 4: Hepatitis C exposure and hepatitis C and HIV testing** |  |  |
| --- | --- | --- |
| **33** | Have you ever had any of the following in the past? (tick all that apply) | 1. Blood transfusion<br>2. Tattoos<br>3. Piercings<br>4. Direct contact with hepatitis C positive blood through a healthcare job (e.g. needlestick injury)<br>5. Medical injections or hospital procedures or surgery (including abortion, endoscopy, dialysis)<br>6. Injections for non-medical purposes (e.g. recreational drug use or traditional healer)<br>7. A close family member having hepatitis C<br>8. None of the above<br>9. Not sure<br>10. Other |
| **34** | Q30 (10) → Please specify | {text} |
| **35** | Has your biological mother ever tested positive for hepatitis C? | 1. No<br>2. Yes<br>3. Don't know |
| **36** | Has a sexual partner or family member of yours ever tested positive for hepatitis C? | 1. No<br>2. Yes<br>3. Don't know |
| **37** | Have <u>you</u> ever had a test for hepatitis C? | 1. No<br>2. Yes<br>3. Don't know |
| **38** | Q37 (2) → When was the last time you were tested? | 1. During this pregnancy<br>2. During a previous pregnancy<br>3. Another time when I was not pregnant |
| **39** | Q37 (2) → What was the result? | 1. Negative<br>2. Positive<br>3. Don't know |
| **40** | Q39 (2) → Do you know if the test result was?<br>(tick all that apply) | **1.** Positive for hepatitis C antibody<br>**2.** Positive for hepatitis C virus (i.e. RNA/PCR positive)<br>**3.** Positive for hepatitis C but not sure if I was tested for antibody or virus (RNA/PCR) |
| **41** | Q37 (2) → Approximately how much did the hepatitis C test cost? | **1.** Nothing, it was free<br>**2.** 100 – 200 UAH<br>**3.** 200 – 300 UAH<br>**4.** 300 – 500 UAH<br>**5.** Don't know<br>**6.** Other |
| **42** | Q37 (6) → Please specify | {text} |
| **43** | Q40 (1-3) → Have you been treated for hepatitis C? | **1.** No<br>**2.** Yes<br>**3.** Don't know |
| **42** | Q41 (2) → How did you take the medicine to treat your hepatitis C? | **1.** Weekly injections plus tablet<br>**2.** Daily tablets only |
| **43** | Q41 (2) → Were you cured? | **1.** No<br>**2.** Yes<br>**3.** Don't know |
| **44** | Q22 (2) & 23 (2) → How many children do you have who are alive today (not including your current/ recent pregnancy)? | {number} |
| **45** | Q44 >0 → Have any of your children been tested for hepatitis C? | **1.** None have been tested<br>**2.** Yes, at least one has<br>**3.** Don't know |
| **46** | Q45 (2) → Did any of your children test positive for hepatitis C? | **1.** No<br>**2.** Yes<br>**3.** Don't know |
| **47** | Q46 (2) → How many tested positive for hepatitis C? | {number} |
| **48** | Do you have HIV? | **1.** No<br>**2.** Yes<br>**3.** Not sure |
| **49** | Q13 (2) and Q48 (2) → Are you currently taking antiretroviral drugs for your HIV (i.e. during your pregnancy)? | **1.** No<br>**2.** Yes |
| **50** | Q13 (1) and Q48 (2) → Are you currently taking antiretroviral drugs for your HIV (i.e. following delivery of your baby)? | **1.** No<br>**2.** Yes |
| **51** | Q48 (2) → Did you take antiretroviral drugs for your HIV during your recent pregnancy? | **1.** No<br>**2.** Yes |
| Section 6: Questions about your possible exposure to Hepatitis C and HIV testing |  |  |
| 52 | At the moment pregnant women in Ukraine have to pay for the test for hepatitis C. Do you think all women should be offered free hepatitis C testing in pregnancy? | 1. No<br>2. Yes<br>3. Don't know |
| 53 | Q52 (1) → Why not? | {text} |
| 54 | **It is not clear whether medicines for curing hepatitis C are safe when taken during pregnancy.**<br>Given this, would you consider taking medicine for hepatitis C during pregnancy? | 1. Yes, to cure me, even if it does not lower the chance of my baby getting hepatitis C<br>2. Yes, but only if it lowers the chance of my baby getting hepatitis C or has other benefits for my baby<br>3. No, I would wait until after my pregnancy to start treatment |
|  | (Please only give one response, whichever you consider your main response) | 4. No, I would wait until there were more studies testing these medicines before I decided to start treatment during pregnancy |
| 55 | **Now, let's pretend that medicines for hepatitis C have been found to be safe for both mother and baby when taken during pregnancy.**<br>Given this, and imagining you are hep C positive, would you consider taking medicine for hepatitis C during pregnancy?<br>(Please only give one response, whichever you consider your main response) | 1. Yes, to cure me, even if it does not lower the chance of my baby getting hepatitis C<br>2. Yes, but only if it lowers the chance of my baby getting hepatitis C or has other potential benefits for my baby<br>3. No, I would wait until after my pregnancy to start treatment |

|  |
| --- |
| Thank you for completing this survey. |
| Please let us know if you have any comments or questions |

## References

1. Dugan E, Blach S, Biondi M, Cai Z, DePaola M, Estes C, et al. Global prevalence of hepatitis C virus in women of childbearing age in 2019: a modelling study. Lancet Gastroenterol Hepatol. 2021;6(3)169–84.

2. Schmelzer J, Dugan E, Blach S, Coleman S, Cai Z, DePaola M, et al. Global prevalence of hepatitis C virus in children in 2018: a modelling study. Lancet Gastroenterol Hepatol. 2020;5(4)374–92.

3. Benhammou V, Tubiana R, Matheron S, Sellier P, Mandelbrot L, Chenadec JL, et al. HBV or HCV Coinfection in HIV-1-Infected Pregnant Women in France: Prevalence and Pregnancy Outcomes. J Acquir Immune Defic Syndr. 2018;77(5)439–50.

4. Malik F, Bailey H, Chan P, Collins IJ, Mozalevskis A, Thorne C, et al. Where are the children in national hepatitis C policies? A global review of national strategic plans and guidelines. JHEP Rep. 2021;3(2)100227.

5. Bhardwaj AM, Mhanna MJ, Abughali NF. Maternal risk factors associated with inadequate testing and loss to follow-up in infants with perinatal hepatitis C virus exposure. J Neonatal Perinatal Med. 2020;10.3233/NPM-190264.

6. Kuncio DE, Newbern EC, Johnson CC, Viner KM. Failure to test and identify perinatally infected children born to Hepatitis C virus-infected women. Clin Infect Dis. 2016;62(8)980–5.

7. Chappell CA, Scarsi KK, Kirby BJ, Suri V, Gaggar A, Bogen DL, et al. Ledipasvir plus sofosbuvir in pregnant women with hepatitis C virus infection: a phase 1 pharmacokinetic study. Lancet Microbe. 2020;1(5)e200–e8.

8. AbdAllah M, Alboraie M, Abdel-Razek W, Hassany M, Ammar I, Kamal E, et al. Pregnancy outcome of anti-HCV direct-acting antivirals: Real-life data from an Egyptian cohort. Liver Int. 2021;41(7)1494–7.

9. Yattoo GN. Treatment of chronic hepatitis C with ledipasvir/sofosbuvir combination during pregnancy. Abstract number O-HCV-38. 27th Annual Conference of Asian Pacific Association for the Study of the Liver; New Delhi, India 2018.

10. Judd A, Collins IJ, Pett S, Gibb DM. Labour pains: eliminating HCV in women and children. Lancet Gastroenterol Hepatol. 2021;6(3)150–1.

11. Kushner T, Cohen J, Tien PC, Terrault NA. Evaluating Women’s Preferences for Hepatitis C Treatment During Pregnancy. Hepatol Commun. 2018;2(11)1306–10.

12. Kouyoumjian SP, Chemaitelly H, Abu-Raddad LJ. Characterizing hepatitis C virus epidemiology in Egypt: systematic reviews, meta-analyses, and meta-regressions. Sci Rep. 2018;8(1)1661–.

13. Al Kanaani Z, Mahmud S, Kouyoumjian SP, Abu-Raddad LJ. The epidemiology of hepatitis C virus in Pakistan: systematic review and meta-analyses. R Soc Open Sci. 2018;5(4)180257.

14. Hope VD, Eramova I, Capurro D, Donoghoe MC. Prevalence and estimation of hepatitis B and C infections in the WHO European Region: a review of data focusing on the countries outside the European Union and the European Free Trade Association. Epidemiol Infect. 2014;142(2)270–86.

15. UNAIDS. Global AIDS Monitoring 2020 Country progress report - Egypt Geneva; 2020.

16. UNAIDS. Global AIDS Monitoring 2020 Country progress report - Pakistan. Geneva; 2020.

17. Hassanin A, Kamel S, Waked I, Fort M. Egypt’s Ambitious Strategy to Eliminate Hepatitis C Virus: A Case Study. Glob Health Sci Pract. 2021;9(1)187–200.

18. Chaillon A, Rand EB, Reau N, Martin NK. Cost-effectiveness of universal hepatitis C virus screening of pregnant women in the United States. Clin Infect Dis. 2019;69(11)1888–95.

19. Deuffic-Burban S, Huneau A, Verleene A, Brouard C, Pillonel J, Le Strat Y, et al. Assessing the cost-effectiveness of hepatitis C screening strategies in France. J Hepatol. 2018;69(4)785–92.

20. Maalej NH, Collins IJ, Ades A, Scott K, Judd A, Chappell E, et al., editors. Modelling the potential effectiveness of different screening and treatment strategies for hepatitis c during pregnancy in Ukraine. International Liver Congress; 2021; Online.

